# Neuropathic Gastroduodenal Disorders can be Diagnosed by Non-Invasive Body Surface Gastric Mapping: a Comparison with Antroduodenal Manometry

**DOI:** 10.1101/2024.11.14.24317342

**Authors:** Christian Sadaka, Binghong Xu, Alain J Benitez, Carolyn M. Orians, Corey Bowerman, Jordan Dourlain, Beth Skaggs, Kimberly G. Konka, Lexi Roshkovan, Gayl Humphrey, Stefan Calder, Gabriel Schamberg, Greg O’Grady, Armen A Gharibans, Hayat Mousa

## Abstract

**Background:** Antroduodenal manometry (ADM) is the gold standard for diagnosing myopathy and neuropathy in patients with upper gastrointestinal (GI) disorders. However, ADM is invasive and costly. Body surface gastric mapping (BSGM) has emerged as a non-invasive test to assess gastric function. This study evaluates the correlation between BSGM and ADM in children.

**Methods:** Clinically indicated ADM was performed using high-resolution water-perfused motility catheters, with simultaneous BSGM. Real-time symptoms were tracked using a validated symptom logging app at 15-min intervals, and Nausea Severity Scale (NSS) scores recorded. Protocols involved a 2 hour fast, provocative testing, a meal, and over 1 hr of postprandial recordings. ADM tracings were categorized into neuropathy, myopathy, postprandial hypomotility, or normal. BSGM metrics included principal gastric frequency, BMI-adjusted amplitude, and Rhythm Index. Gastric emptying data were also evaluated.

**Results:** Fifteen subjects were recruited (13 female; ages 10-19 years, BMI 20.6±3.5 kg/m^2^). ADM results showed neuropathy (n=4), myopathy (n=1), post-prandial hypomotility (n=4), and normal motility (n=6). Patients with neuropathy had lower rhythm stability (0.16±0.03 vs 0.38±0.17; p=.001), accompanied by higher nausea scores (8.2±1.2 vs 3.15±2.9; p<.001) and bloating (7.3±0.9 vs 2.1±2.0; p<.001). ADM neuropathy correlated completely with dysrhythmia on BSGM. Normal ADM were consistent with normal BSGM in 83.3% of cases. Delayed gastric emptying did not correlate with a neuropathic diagnosis.

**Conclusion:** Dysrhythmic BSGM phenotype reliably identified gastrointestinal neuropathy with identical results to ADM, with significant correlations to nausea and bloating severity. This study supports considering BSGM as an actionable biomarker when performing ADM is not feasible.

**Key points:**

– Body Surface Gastric Mapping (BSGM) low stability phenotype correlated perfectly with neuropathy detected on Antroduodenal Manometry (ADM). The BSGM normal phenotype also showed a good correlation with normal ADM studies.
– BSGM low stability phenotype was linked to higher severity scores for nausea and bloating.

Abbreviations used: Antroduodenal Manometry (ADM), Body Surface Gastric Mapping (BSGM), Gastric Emptying Scintigraphy (GES), Gastroparesis (GP), Functional Dyspepsia (FD), Electrogastrography (EGG)

## Introduction

Gastroparesis (GP) and functional dyspepsia (FD) are common pediatric gastroduodenal disorders that present a diagnostic challenge due to the significant overlap in their clinical presentations ^(1)^. This overlap includes common symptoms such as early satiety and postprandial fullness, nausea, vomiting, abdominal pain, and bloating ^(2)^. The current practice is to differentiate these two conditions based on gastric emptying (GE) rate in the absence of any mechanical obstruction ^(1)^.

The most commonly used diagnostic tool for differentiating GP and FD is gastric emptying scintigraphy (GES) ^(3)^. However, GES results are not always reliable due to high variability. A recent study reported that one year after an initial GES, 42% of GP patients and 37% of FD patients had results that placed them in the opposite category based on emptying results alone ^(4)^. Moreover, delayed GES is poorly correlated with symptom severity ^(5)^.

Gastric physiology depends on an intact gastric pacemaker, enteric nervous system (ENS), autonomic system, and bidirectional communication between the ENS and central nervous system (CNS), encompassed within the gut-brain axis ^(6, 7)^. Previously, electrogastrography (EGG) was attempted to measure the output of gastric motility mechanisms non-invasively; however, its clinical utility was limited use due to weak clinical correlations ^(8)^. Other limitations included signal loss, high susceptibility to artifacts, lack of spatial analysis, and inconsistent methodology ^(8, 9)^. Currently, the gold standard to confirm the integrity of the ENS in patients with severe gastric symptoms is antroduodenal manometry (ADM). ADM can distinguish between neuropathy, myopathy, and to diagnose post-prandial hypomotility, rumination, pylorospasm, visceral hyperalgesia, and confirm or exclude pseudo-obstruction ^(10)^. However, ADM is an invasive and cost-intensive procedure that is only currently available in major specialist centers.

In recent years, a new generation of multi-channel high-resolution technology has been introduced to advance non-invasive gastric diagnostics, called Body Surface Gastric Mapping (BSGM) ^(11)^. BSGM is a new FDA approved diagnostic tool that measures gastric myoelectrical activity and associated contractions using an array of 64 electrodes connected to a wearable reader, with simultaneous validated symptom profiling ^(12, 13)^. The outputs include a graphical representation and metrics of the gastric frequency, BMI-adjusted amplitude, rhythm, and meal response ^(9, 14)^. Several studies have been published in adults and pediatrics with promising results on patient phenotyping and symptom correlations, including those with Nausea and Vomiting Syndrome (NVS), long-term diabetes, and adolescents with gastroparesis and functional dyspepsia ^(15–18)^.

The aim of this study was to assess the degree of correlation between ADM motility patterns and BSGM phenotypes in children. We hypothesized that different BSGM phenotypes would correlate with specific ADM findings, and that BSGM could potentially serve as an actionable biomarker comparable to ADM. We also hypothesized that BSGM would be able to define specific underlying mechanisms of symptoms, whether related to the pacemaker, neuropathy, or other causes. Gastric emptying data were also evaluated.

## Materials and Methods

This study was a prospective single center study, in which patients scheduled for a clinically indicated ADM consented to have BSGM conducted simultaneously. The BSGM has been approved at the Children’s Hospital of Philadelphia (CHOP) since 2021. The IRB approved BSGM protocol modifications to accommodate the clinically indicated and approved ADM protocol.

### ADM Protocol

The ADM protocol at the Children’s Hospital of Philadelphia is based on the ANMS-NASPGHAN consensus ^(10)^. The ADM catheter was placed by an Interventional Radiology attending under sedation. Manometry was performed using a water perfused motility catheter, with placement guided by fluoroscopy, consisting of 16 channels spaced at three-centimeter intervals. The ADM protocol included a two-hour fasting period, followed by an hour of post-erythromycin infusion observation. A second erythromycin infusion with a higher dose was administered at the clinician discretion, based on the response to first erythromycin dose. Erythromycin was given to induce a phase III Migratory Motor Complex (MMC) in the antrum and intestine ^(10)^. Patients selected a full portion age-appropriate meal to be consumed over a maximum of 30 minutes; calorific value varied with age and meal selection. Patients were encouraged to try to complete the meal. Patients are monitored for 2 hours post-prandially followed by the administration of octreotide and 30 minutes of monitoring. Octreotide was administered to induce phase III MMC activity in the small bowel with antral inhibition ^(10)^.

### BSGM Protocol

BSGM recordings were performed using the Gastric Alimetry® device (Alimetry, New Zealand). After placement of the ADM catheter and the start of motility study, a member of the research team placed the BSGM array on patient’s abdomen, with the exact placement calculated according to a built-in algorithm to localize the antrum and pylorus. Placement of the array also followed a standardized protocol of skin preparation, and signal processing ^(11)^. We then connected the BSGM array to a reader which was also linked to an iPad by Bluetooth. BSGM recordings commenced after one and a half hour of fasting, which allowed capture of a minimum of thirty minutes of fasting activity after excluding the 1-hour post-prokinetic phase.

Figure 1 shows the study protocol including ADM and BSGM.

**Figure 1:** Panel A: Antroduodenal manometry Protocol used at the Children’s Hospital of Philadelphia Panel B: Standard Body Surface Gastric Mapping Protocol Panel C: Antroduodenal Manometry-Body Surface Gastric Mapping Study Protocol

### Symptom assessment

Patients completed the Nausea Severity Score questionnaire to assess their symptoms experienced over the past 2 weeks ^(19)^. In addition, throughout the study, patients continuously recorded their symptoms (nausea, abdominal pain, excessive fullness, bloating, heartburn, and stomach burn) along with any episodic symptom (vomiting, reflux, belching) using a validated app ^(13)^.

### Patient Recruitment

Patients aged between 8 to 25 years who were scheduled for a clinically indicated ADM were deemed suitable for inclusion. Exclusion criteria included pregnancy (tested in age-appropriate females), significant cognitive impairment, body mass index of >35, active abrasions or inflammations on abdominal skin, history of skin allergies, severe allergies to adhesives, cosmetics, or lotions, inability to fast for 6 hours, or inability to maintain a quiet reclined position for 4.5 hours. Parental/guardian permission (informed consent) and if appropriate, child assent was obtained from all patients.

We reviewed all patients’ charts on EPIC. Diagnoses and clinical information were obtained from their charts. All patients also had prior GES within a year before the ADM. Gastric emptying values of 40% at 2 hours and 90% at 4 hours were used as normal reference values for GES ^(20)^. Dysautonomia diagnosis was based on clinical orthostatic changes; no objective testing was done to confirm dysautonomia in these patients.

### Analysis

Upon analyzing the ADM tracing, we identified five metrics: motility index, number of peaks, peaks/minute, mean amplitude of peaks, and the area under the curve. These metrics along with the tracings were used to define four diagnostic ADM tracing patterns: normal, neuropathic dysmotility, post prandial antral hypomotility, and myopathic dysmotility. The interpretation was performed based on ANMS-NASPGHAN consensus and GLASS scoring ^(10, 21)^. ADM tracing analysis was independently performed by two motility experts without reference to the BSGM recordings, and any differences were settled by consensus.

BSGM data were analyzed using the automatically generated FDA-cleared Gastric Alimetry Report v3.1.0 (Alimetry Limited, Auckland NZ) ^(22)^. Four study metrics were assessed: principal gastric frequency, Rhythm Index, BMI-adjusted amplitude, and Fed:Fasted amplitude ratio ^(14)^.The principal gastric frequency represents the average frequency in cycles per minute (cpm), in which the electrical signals were being recorded while the Rhythm Index describes the stability of the gastric activity with its concentration around the principal gastric frequency; the BMI adjusted amplitude describes the amplitude of the gastric frequency after adjustments based on BMI; and the Fed:Fasted ratio compares the amplitude post prandially to that in the fasting period ^(14)^. The reference values for these metrics in pediatrics are shown in Supplementary Material 1 ^(23)^. In addition, a total symptom burden score is calculated using the scores from each reported symptom ^(14)^.

Statistical analyses were performed using SPSS version 29.0 (IBM Corporation, Armonk, NY). Variables were summarized with descriptive statistics, either as mean with standard deviation (SD) or median and interquartile range (IQR). Statistical comparisons were performed between study groups using Student’s t-test, chi-squared tests, Mann-Whitney U, Kruskal-Wallis, or Fisher’s exact tests as appropriate.

## Results

### Patient Demographics

A total of 15 patients underwent simultaneous ADM and BSGM. The majority of patients were female (13/15; 87%), and the mean age was 15.9 ± 2.7 (range 10-19 years). The mean BMI was 20.6 ± 3.5. Eleven patients had the catheter placed intranasally (73.33%), while the other 4 had the catheter placed through an existing Gastrostomy-tube (26.67%). At enrollment, 8 patients had a diagnosis of GP, while 7 were diagnosed with FD. Patient demographics are summarized in Supplementary Material 2.

Referral indications for ADM included abdominal pain in 12 patients, nausea in 12 patients, and suspected motility disorder in 11 (see supplementary material 3 for all the clinical indications). The most common comorbid conditions were dysautonomia (n=8), a psychological condition (n=6), and asthma (n=5) (refer to Figure 2). Nine patients were prescribed prokinetics, five patients PPIs, four SSRIs, and three cyproheptadine. Prokinetics and cyproheptadine were discontinued at least 2 days prior to the study.

**Figure 2:**
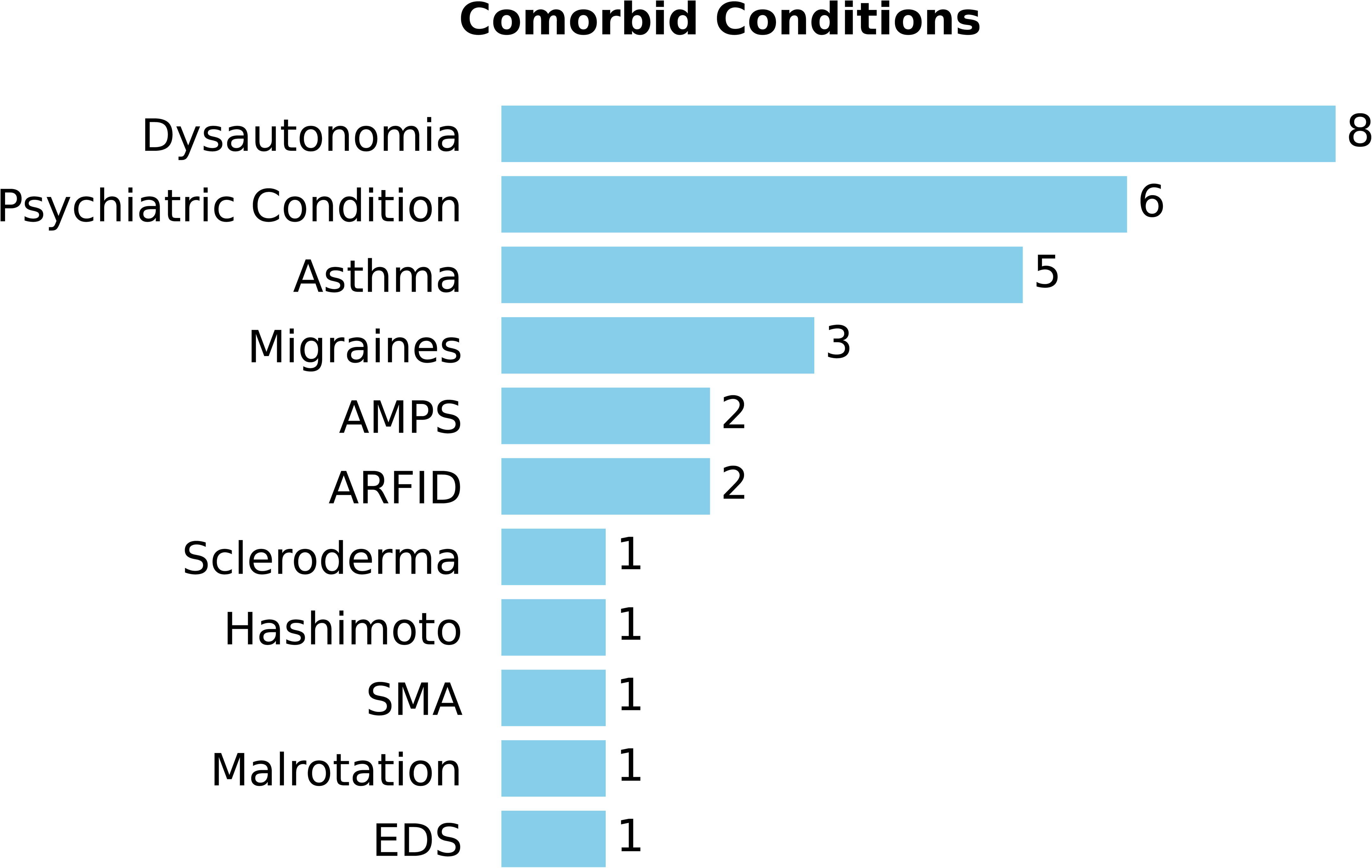
Patient comorbid conditions

### ADM Tracings and BSGM Phenotypes

Figure 3 shows examples of four different classes of ADM tracings recorded in this study and Figure 4 shows the three different BSGM phenotypes recorded. Per the ANMS-NASPGHAN consensus and GLASS scoring ^(10, 21)^, four out of the 15 patients were diagnosed with neuropathic dysmotility, another four had post prandial antral hypomotility, one patient had myopathic dysmotility, while the remaining six had normal studies. BSGM reports identified four patients with a low stability phenotype, evidenced by a Rhythm Index <0.25, four patients had a low Fed:Fasted amplitude ratio on BSGM, and eight patients had a normal BSGM ^(17)^.

**Figure 3:**
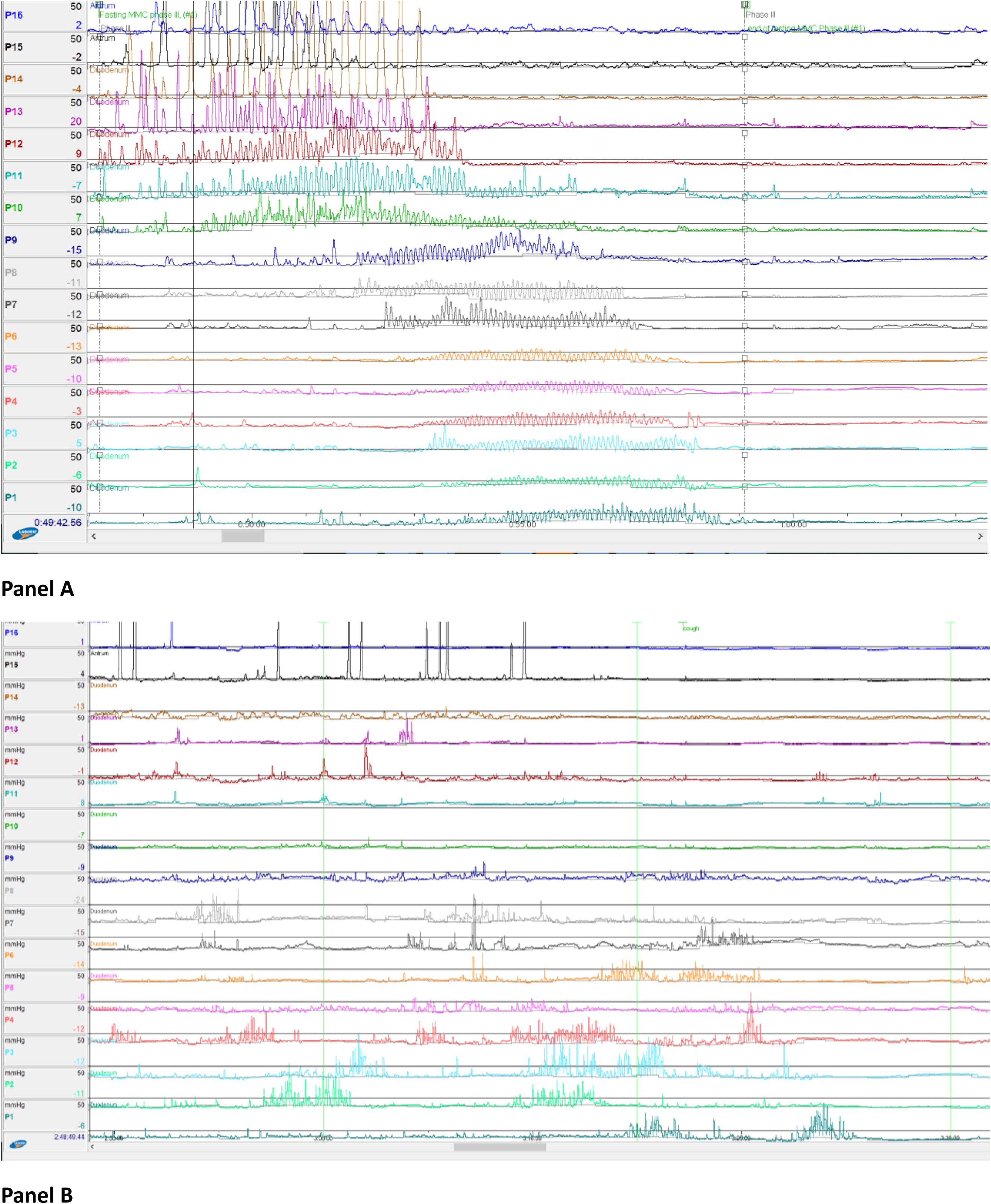
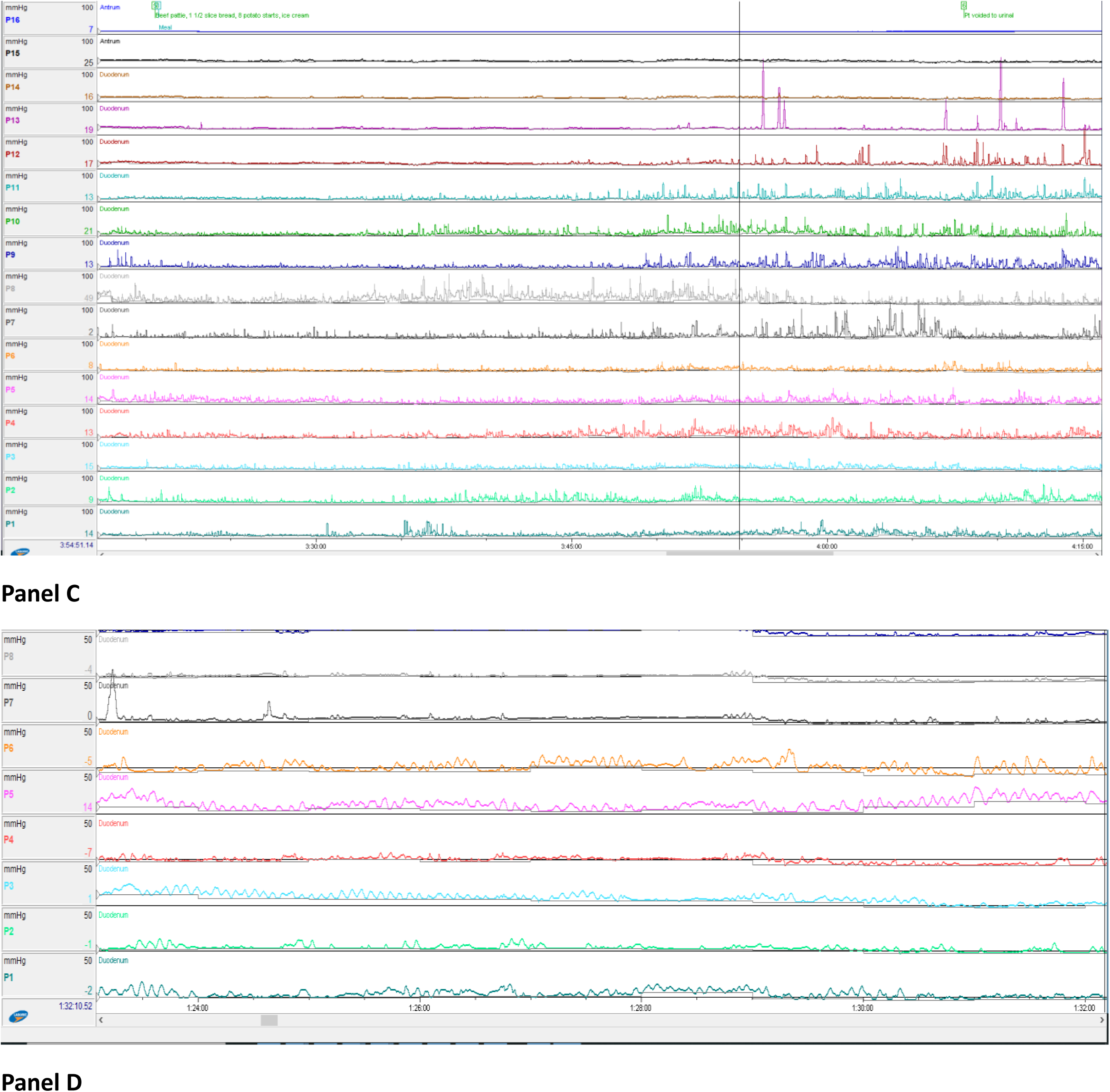
Panel A: Normal antroduodenal manometry tracing Panel B: Neuropathic antroduodenal manometry tracing Panel C: Post-Prandial Antral Hypomotility antroduodenal manometry tracing Panel D: Myopathic antroduodenal manometry tracing

**Figure 4:**
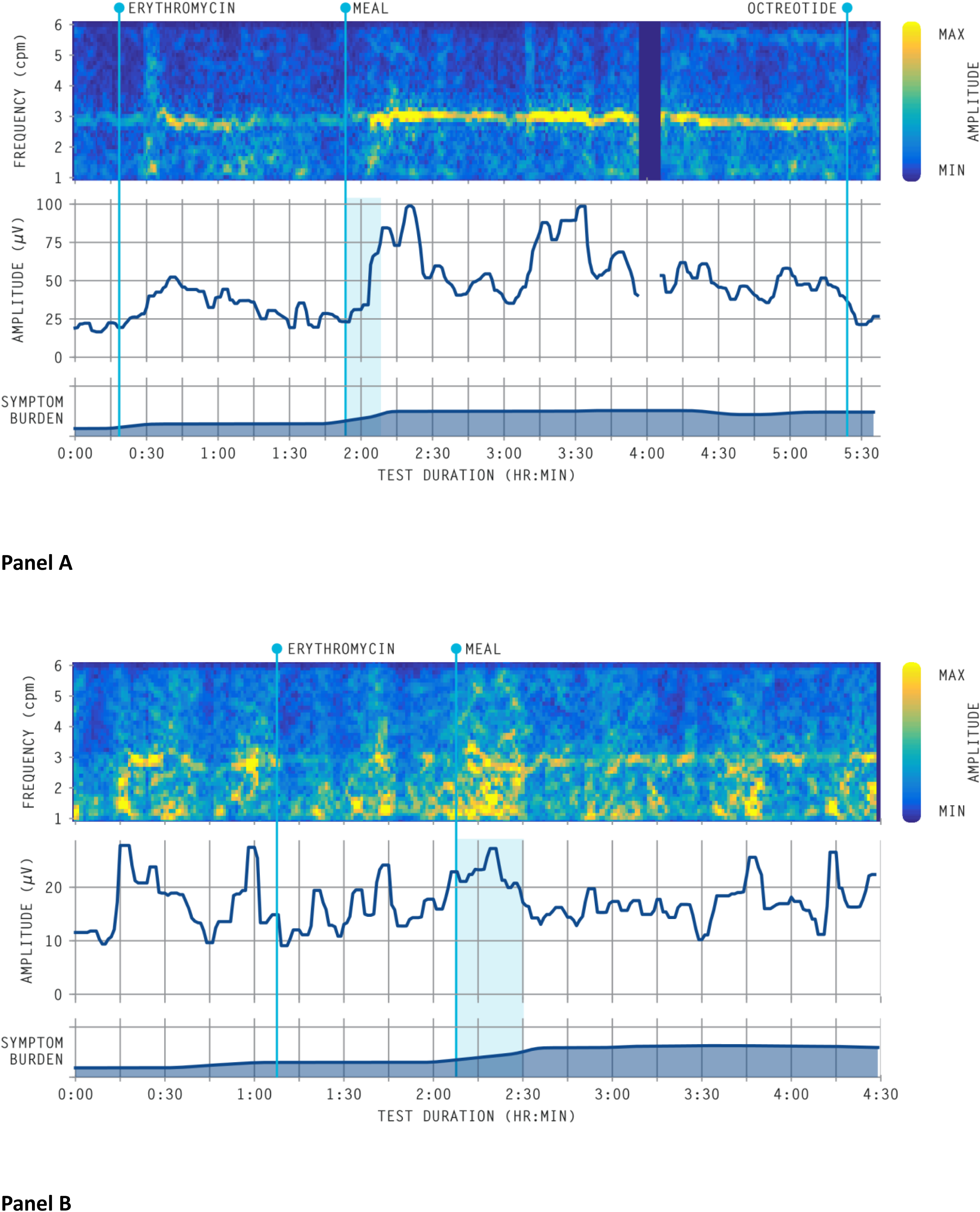
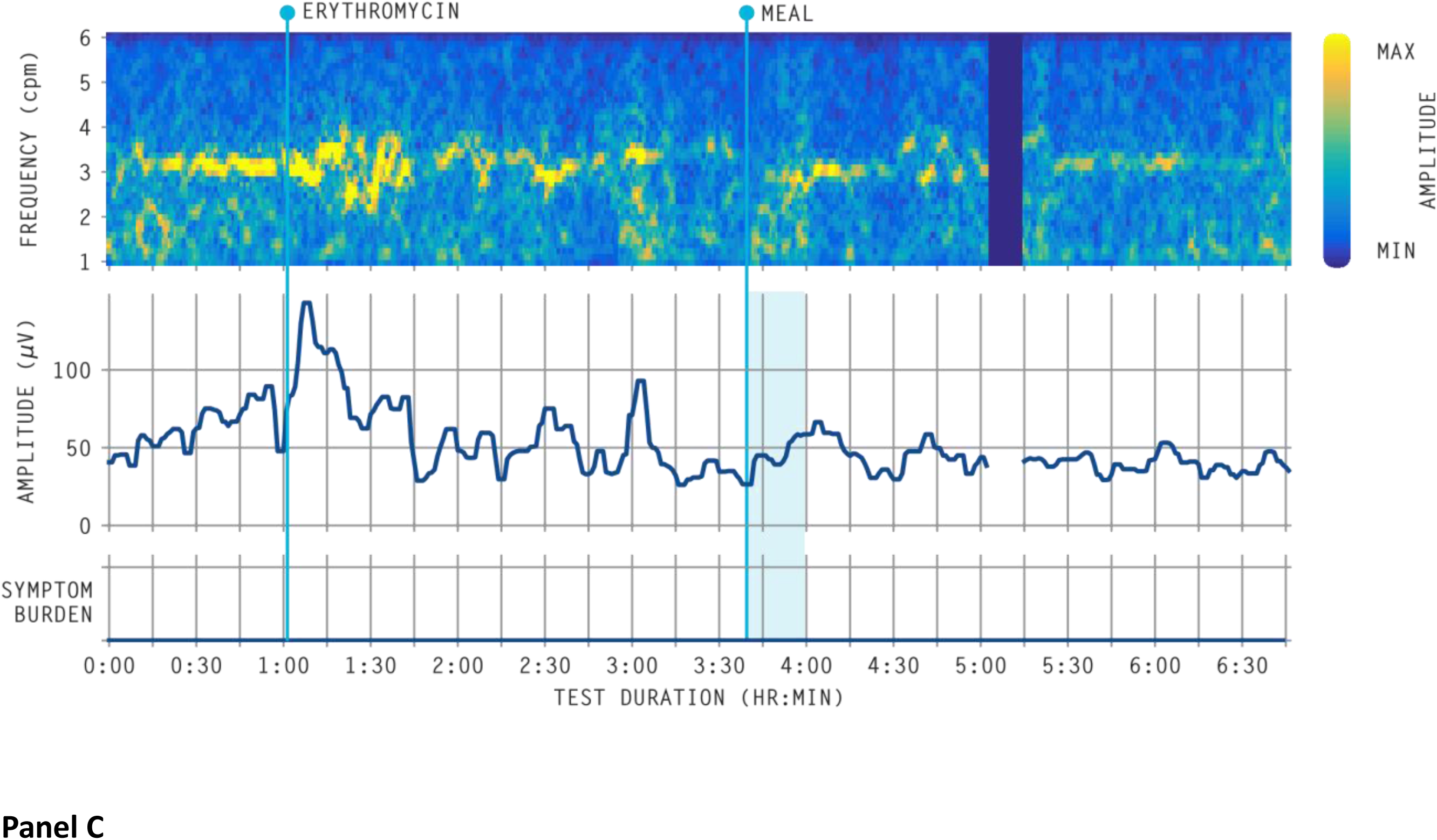
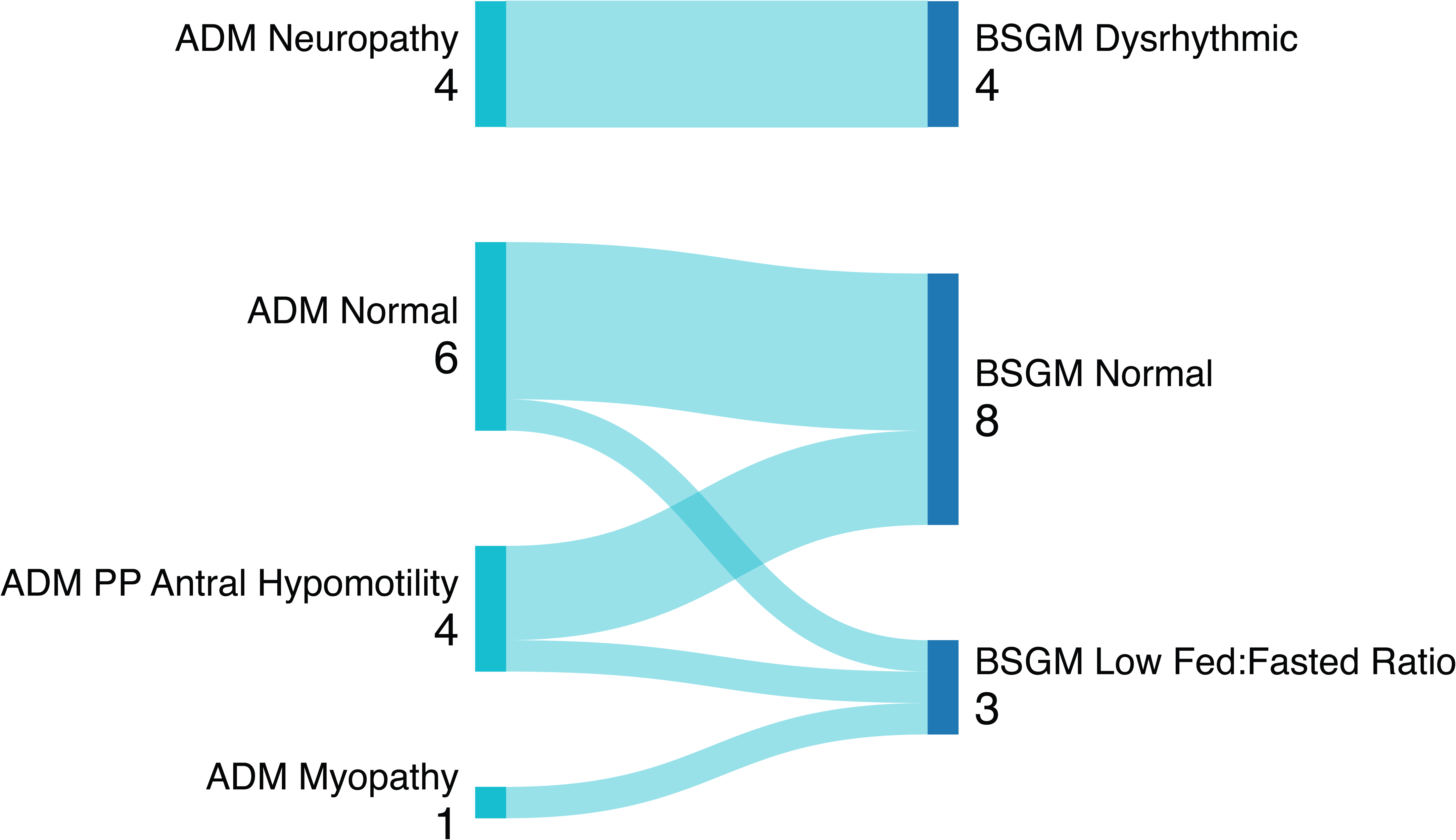
Panel A: Normal Body Surface Gastric Mapping phenotype graph Panel B: Low Stability Body Surface Gastric Mapping phenotype graph Panel C: Low Fed:Fasted Body Surface Gastric Mapping phenotype graph

Figure 5 presents the comparison between ADM and BSGM results. All four patients with neuropathic ADM had a low stability BSGM phenotype, indicating a complete correlation. In addition, the Rhythm Index was significantly lower in the neuropathic group compared to the non-neuropathic patients (0.16 ± 0.03 vs 0.38 ± 0.17) (p=0.001). Of the six patients with normal ADM studies, five had normal BSGM, while one had a low Fed:Fasted ratio. The myopathy patient also had a low Fed:Fasted ratio. Of the four post-prandial antral hypomotility patients, one had a low Fed:Fasted ratio, and three patients had a normal BSGM phenotype.

**Figure 5:**
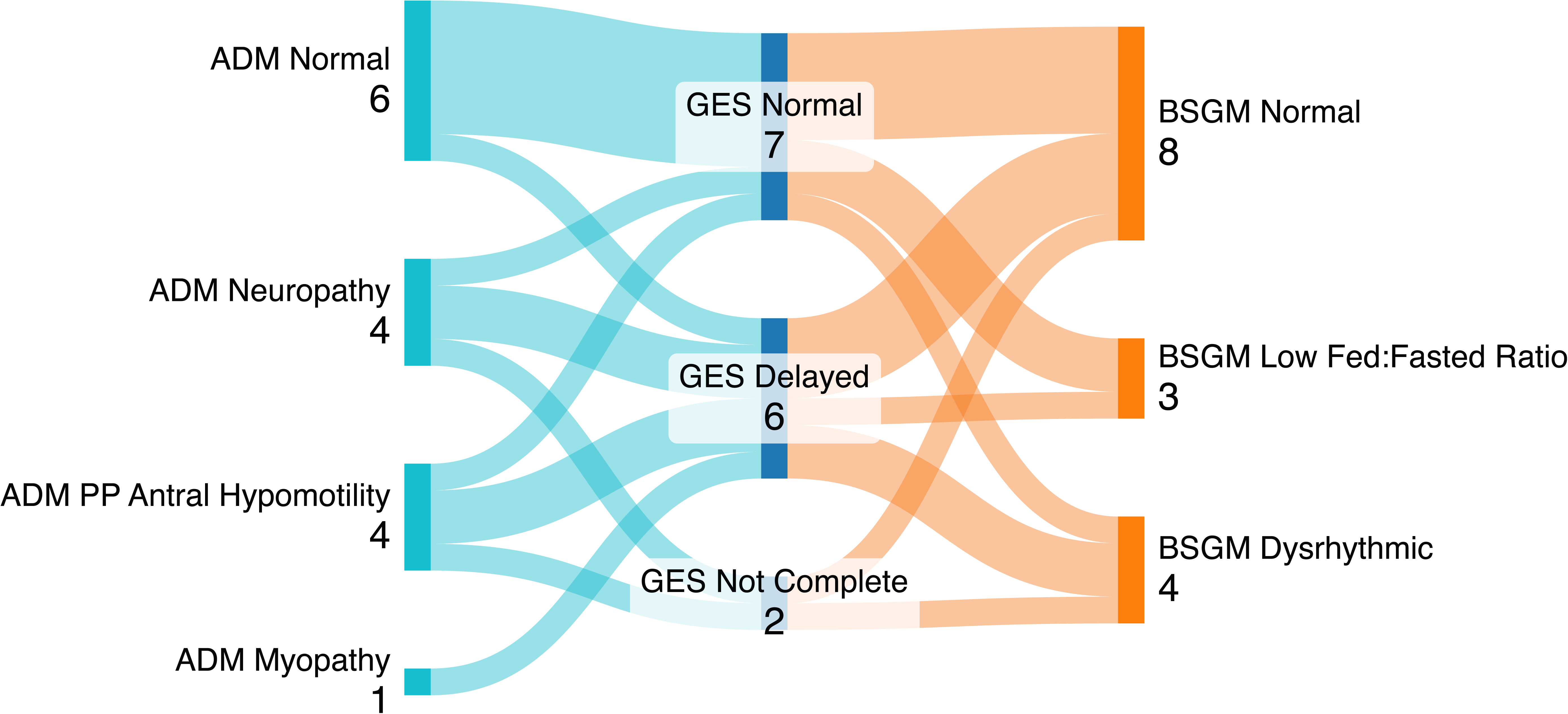
Sankey Diagram showing the correlation between Antroduodenal manometry results and Body Surface Gastric Mapping phenotype

Of the eight patients reported with dysautonomia, four had neuropathy on ADM and low stability on BSGM. Of the remaining four, one had a normal ADM, while three had post-prandial antral hypomotility. BSGM testing found that two of these four patients had a normal phenotype while two had a low Fed:Fasted ratio.

### Patient Symptomatology

There was no statistical difference in patients’ nausea severity when comparing their 2-week recall Nausea Severity Score between patients whether grouped according to their ADM diagnosis, BSGM phenotype, or GES result. During the BSGM study, all subjects reported their real time symptoms (nausea, abdominal pain, excessive fullness, bloating, heartburn, and stomach burn) every 15 minutes, and entered the scores on the Gastric Alimetry App (Alimetry Limited, Auckland, NZ). These data showed that the low stability BSGM phenotype was highly correlated with more severe nausea scores (8.2 ± 1.2 vs 3.2 ± 2.9) (p<0.001), being more severe than all other patients (Figure 6A). Low stability BSGM patients also reported higher bloating scores (7.3 ± 0.9 vs 2.1 ± 2.0) (p<0.001) compared to other patients (Figure 6B). The total symptom burden score was also higher in the low stability phenotype (31.4 ± 8.7 vs 21.9 ± 16.4) compared to Normal and fed to fasted Phenotype, however this difference was not statistically significant (p=0.18) (Figure 6C). There was also no statistical difference in the remaining four real-time symptoms reported.

**Figure 6:**
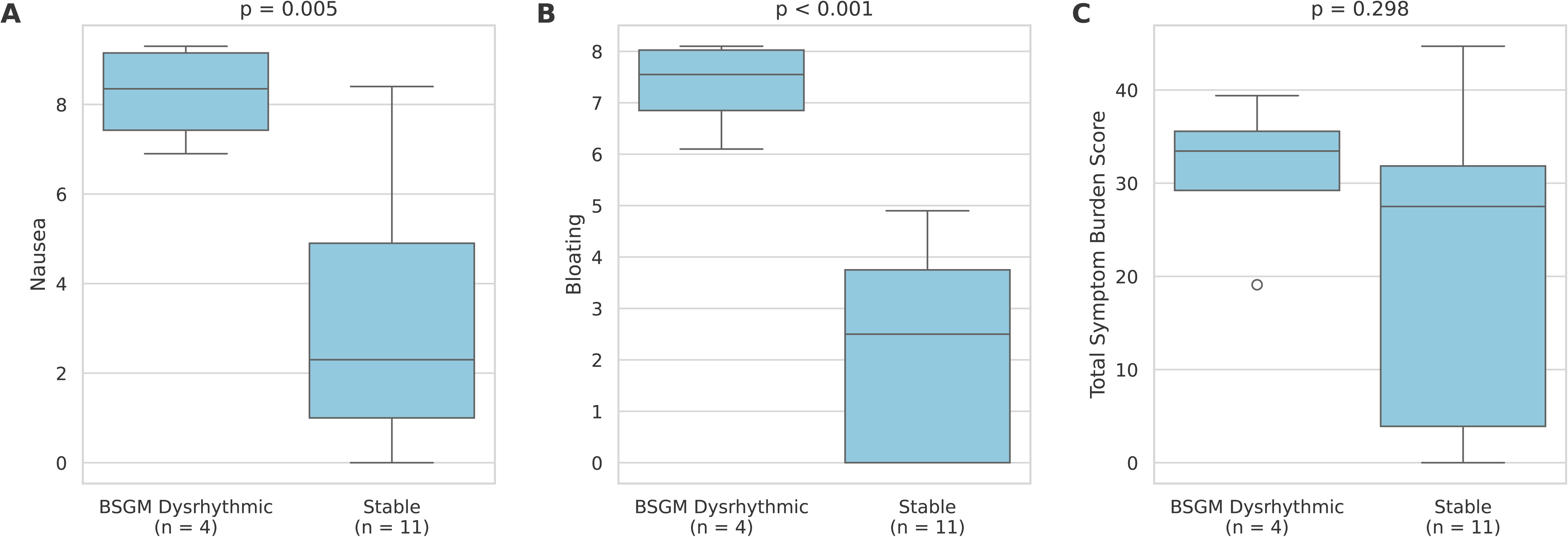
Panel A: Nausea scores comparison between low stability phenotype and normal and low Fed:Fasted phenotype Panel B: Bloating scores comparison between low Stability phenotype and Normal and low Fed:Fasted phenotype Panel C: Total symptoms burden scores comparison between low stability phenotype and Normal and low Fed:Fasted phenotype

### Gastric Emptying

Thirteen patients had a completed GES; two patients were unable to complete the GES and therefore are excluded from this analysis. The majority of GES in this cohort lasted two hours as the two hours emptying result was either abnormal or high in the normal range, for example 70%.

Nine patients had completed more than one GES study. For these patients, we used the most recent to compare GES results to ADM and BSGM results. As shown in Figures 7A and 7B, there was an overall poor correlation between GES and ADM, or GES and BSGM.

**Figure 7:**
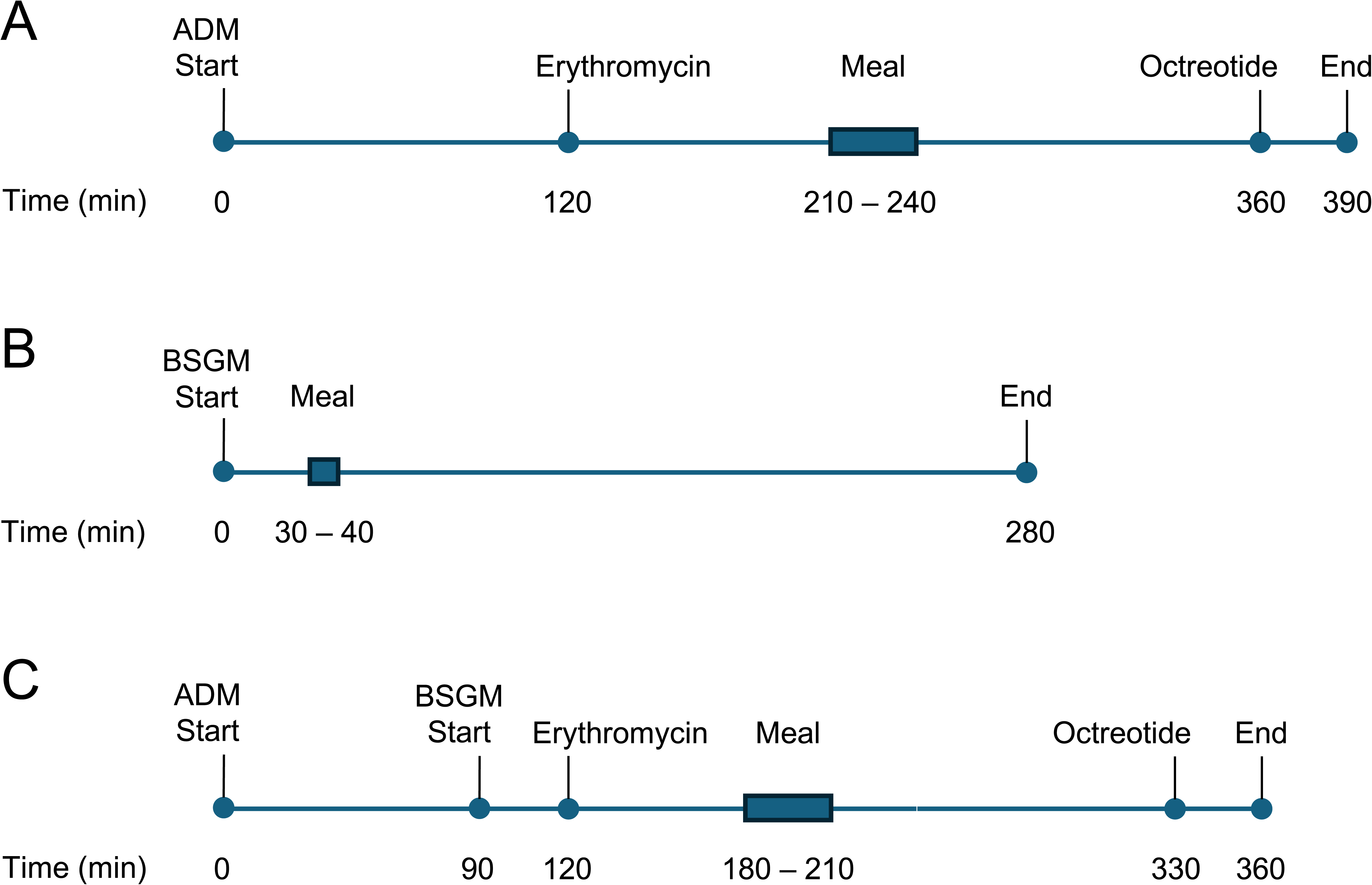
Panel A: Sankey Diagram showing correlation between Antroduodenal manometry and Gastric Emptying Scan Panel B: Sankey Diagram showing correlation between Body Surface Gastric Mapping and Gastric Emptying Scan

As a further analysis, GES outcomes were also compared across time for those nine patients who had multiple studies completed. Initial GES showed delayed gastric emptying in six patients while three had a normal study. GES result normalized in four out of the six patients initially diagnosed with gastroparesis (66.7% change) and two remained delayed. One of the three patients with a normal GES initially developed delayed gastric emptying (33% change) while two patients’ studies remained normal. Hence, out of the nine patients with multiple GES studies, five of them showed clear inconsistency in the result of their GES (55.6%), and a reproducibility of 44.4% only for the evaluated protocol. Figure 8 shows the results of clinically indicated repeat GES on nine patients. Since most studies lasted only 2 hours, we reported the rate of reproducibility at 2 hours emptying in all the patients in the figure.

**Figure 8:**
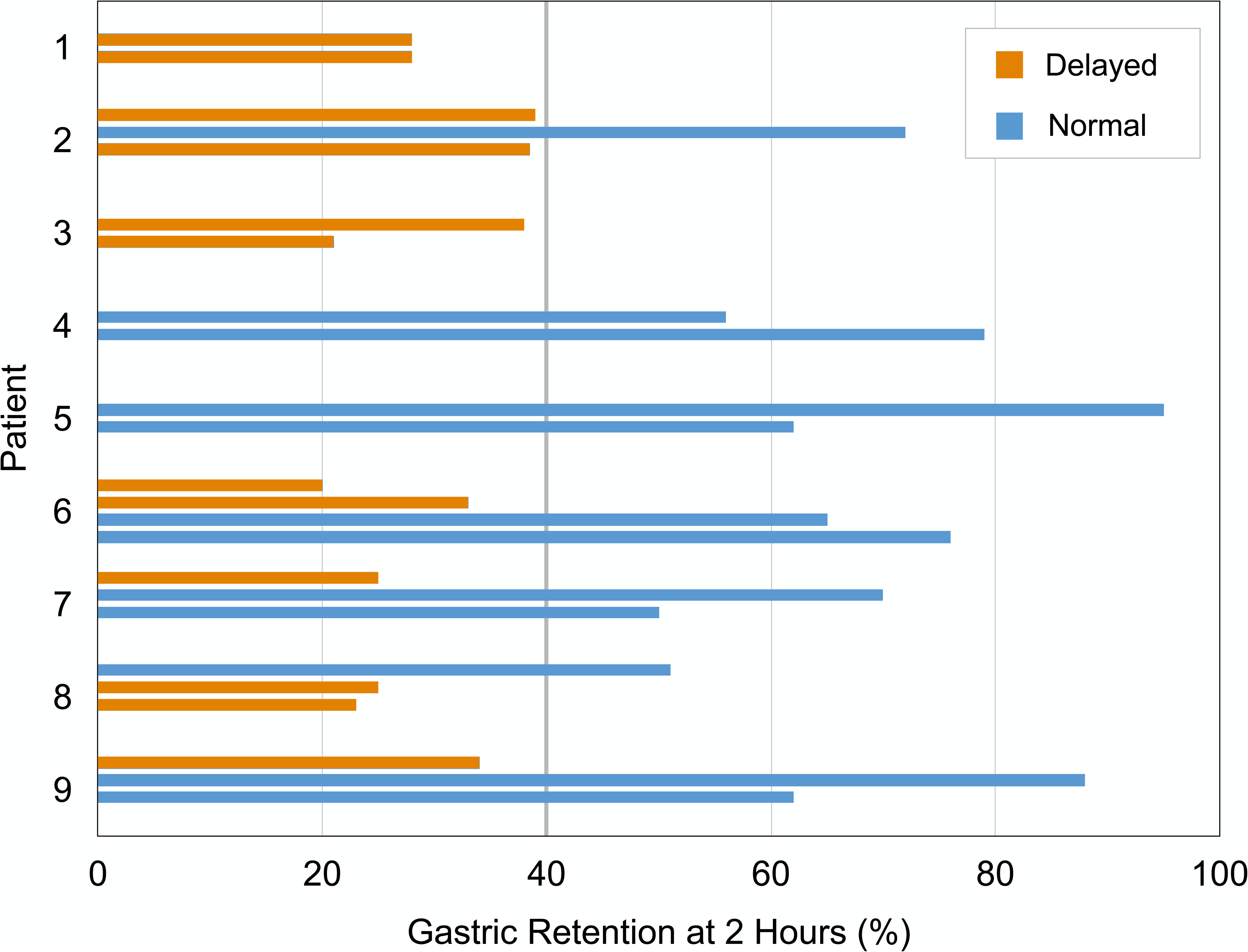
Gastric emptying scan results of patients with multiple studies. Each cluster of studies indicates repeated gastric emptying scans from the same patient.

## Discussion

This is the first study comparing simultaneous gastric myoelectrical activity detected by BSGM to manometric tracing detected by ADM. The key finding was of a close concordance between a neuropathic diagnosis on ADM and the dysrhythmic BSGM phenotype, indicated by a low Rhythm Index, which was also associated with more severe patient symptoms, (nausea and bloating). Moreover, we found that a normal BSGM phenotype was highly correlated with a normal ADM, except for one case of post-prandial antral hypomotility. These results demonstrate a major advance in our understanding of BSGM as an emerging diagnostic tool for identifying specific disease mechanism in patients with upper gut symptoms, associating symptoms with certain BSGM phenotypes, confirming the integrity of gastric biologic pacemaker, and detecting the presence of neuropathy.

An important finding of this study was that the low stability BSGM phenotype was significantly associated with more severe nausea and bloating. Even though some patients with normal ADM had high levels of nausea, low stability was still associated with substantially higher levels of nausea (p=<0.001). Notably, these results confirm the findings from another pediatric BSGM study we recently performed in 25 adolescents with functional dyspepsia and gastroparesis ^(17)^. In that study, the low stability phenotype was likewise significantly associated with higher nausea, bloating and pain scores, quality of life, and a worse functional disability index (FDIs). Meanwhile, the results of gastric emptying did not correlate with any of these symptoms or physical health metrics. The converging data between these two studies reinforces the emerging concept that patients with dysrhythmic BSGM phenotype have more severe symptoms and would typically require more intensive medical management ^(15)^. The ability of BSGM to differentiate neuropathic patients and correlate symptomatology with phenotype thus has clinical importance and indicates a superior diagnostic value compared to GES especially in pediatrics.

Gastric emptying testing in adults is known to have low reproducibility over time and offers suboptimal sensitivity and specificity in identifying neuromuscular disorders ^(4, 5)^. These findings are consistent with our pediatric population, as 55.6% of patients with multiple GES studies showed a clear switch between normal gastric emptying and an emptying that meets current definition of gastroparesis. Although a 2-hour GES test was standard in this study, the values are nevertheless comparable to adult data reported by Parischa et al using a standard 4-hr test^(4)^. By contrast, BSGM was found to be reproducible by Law et al. over both the short and long terms as seen in symptomatic and control adults ^(24)^, thereby providing clinicians with greater assurance regarding their management decisions when based on BSGM phenotypes.

Per adult studies, a low Rhythm Index is considered to be a biomarker of neuromuscular disorders, likely reflecting underlying interstitial cell of Cajal (ICC) loss and injury in the majority of cases ^(15, 25)^. ICC play an integral role in gastric and small intestine motility, as they act as the pacemakers, generating and propagating slow waves to coordinate motility patterns ^(7)^. ICC depletion has been identified as a dominant pathology in subsets of patients with both chronic nausea and vomiting syndrome and gastroparesis, and these patients also have a high rate of gastric dysrhythmias ^(15, 25–27)^. In addition, the NIH Gastroparesis Clinical Research Consortium recently reported similar histological findings of ICC depletion in patients with severe functional dyspepsia ^(28)^. The concordance between the neuropathic phenotype on ADM and dysrhythmic BSGM phenotype in this study now further supports the role of BSGM in reliably detecting such GI neuromuscular abnormalities non-invasively. Interestingly, manometry studies of 88 patients with gastroparesis showed abnormalities suggestive of neurogenic derangement in the proximal small bowel, especially in patients with documented delayed gastric emptying ^(29)^. These data confirm earlier observations of small intestinal involvement in the neuropathic process in some patients who present with upper GI symptoms suggestive of GP ^(30)^.

The relationship between neuropathic ADM and low rhythm stability on BSGM warrants further exploration, however, given that ADM assesses antroduodenal pressure waves whereas BSGM evaluates gastric myoelectrical activity. One explanation for this concordance is that the pathophysiology impacting ICC, most commonly involving abnormal immune activation in idiopathic cases, are likely to be operating across the GI tract. Furthermore, ICC deficits also frequently co-exist with myenteric inflammatory infiltrate and are tightly co-regulated with enteric nervous system function (which may be likewise impaired),meaning that neuropathic ADM and dysrhythmic BSGM share a common underlying constellation of pathological features indicative of true neuromuscular disorders ^(31, 32)^. It is also notable that the dysrhythmic phenotype was detected during fasting, as well as post-prandially.

Generalized gastrointestinal dysmotility presenting with nausea, bloating, early satiety, dysphagia, constipation, diarrhea, and abdominal pain can be a manifestation of dysautonomia ^(33)^. Autonomic dysfunction has been shown to result in delayed transit time in the foregut with delayed gastric emptying being the most common transit abnormality in a cohort of idiopathic autonomic dysfunction patients ^(34)^. Indeed, GI symptoms are frequently seen in Postural Orthostatic Tachycardia Syndrome (POTS), a form of dysautonomia, with abnormal gastric emptying being a common feature ^(35)^. As mentioned above, eight of our patients had a diagnosis of dysautonomia with four of them having clear neuropathic changes on ADM, three of them with antral hypomotility, and only one normal.

This study has several limitations. The primary limitation is the modification of the standard BSGM protocol to align with the clinical ADM protocol which resulted in a reduced postprandial period of 1.5-2 hours instead of the standard four hours, variability in patient meals (with some not meeting the caloric target) and the addition of erythromycin and or octreotide as indicated by the ADM protocol ^(9, 14)^. These factors could have influenced the BSGM meal response and affected the accuracy of the Fed / fasting amplitude ratio ^(36)^. Additionally, the small sample size of 15 patients reflects the challenges inherent in conducting intensive research within a vulnerable patient population. It is nevertheless highly notable that a complete neuropathic case concordance and highly significant endpoints were realized despite these factors.

In conclusion, this study is the first to report simultaneous ADM and BSGM, finding a close concordance between BSGM phenotype and ADM diagnosis. In particular, the low stability phenotype on BSGM matches neuropathic antroduodenal dysmotility on ADM and correlates with worse symptoms. The low stability BSGM phenotype reflects a more severe patient phenotype, which is not detectable by GES.

## Data Availability

All data produced in the present study are available upon reasonable request to the authors

## Acknowledgements

We would like to thank the participants and their families for accepting to participate in our study. We are also grateful to the Children’s Hospital of Philadelphia Frontier Program, grant (Center for Precision Diagnosis and Therapy for Pediatric Motility Disorders) who financially supported the study. We also thank the CHOP endoscopy suite nurses for their help during all the studies.

## References

1. Sato H, Grover M. Gastroparesis and Functional Dyspepsia: Spectrum of Gastroduodenal Neuromuscular Disorders or Unique Entities? Gastro Hep Adv. 2023;2(3):438–48.

2. Lacy BE, Tack J, Gyawali CP. AGA Clinical Practice Update on Management of Medically Refractory Gastroparesis: Expert Review. Clin Gastroenterol Hepatol. 2022;20(3):491–500.

3. Camilleri M, Kuo B, Nguyen L, Vaughn VM, Petrey J, Greer K, et al. ACG Clinical Guideline: Gastroparesis. Am J Gastroenterol. 2022;117(8):1197–220.

4. Pasricha PJ, Grover M, Yates KP, Abell TL, Bernard CE, Koch KL, et al. Functional Dyspepsia and Gastroparesis in Tertiary Care are Interchangeable Syndromes With Common Clinical and Pathologic Features. Gastroenterology. 2021;160(6):2006–17.

5. Carbone F, De Buysscher R, Van den Houte K, Schol J, Goelen N, Tack J. Relationship Between Gastric Emptying Rate and Simultaneously Assessed Symptoms in Functional Dyspepsia. Clin Gastroenterol Hepatol. 2022;20(3):e429–e37.

6. Corsello A, Scatigno L, Govoni A, Zuccotti G, Gottrand F, Romano C, et al. Gut dysmotility in children with neurological impairment: the nutritional management. Front Neurol. 2023;14:1200101.

7. Foong D, Zhou J, Zarrouk A, Ho V, O’Connor MD. Understanding the Biology of Human Interstitial Cells of Cajal in Gastrointestinal Motility. Int J Mol Sci. 2020;21(12).

8. Bhat S, Varghese C, Carson DA, Hayes TCL, Andrews CN, Mousa H, et al. Electrogastrography Abnormalities in Pediatric Gastroduodenal Disorders: A Systematic Review and Meta-analysis. J Pediatr Gastroenterol Nutr. 2021;73(1):9–16.

9. O’Grady G, Varghese C, Schamberg G, Calder S, Du P, Xu W, et al. Principles and clinical methods of body surface gastric mapping: Technical review. Neurogastroenterol Motil. 2023;35(10):e14556.

10. Rosen R, Garza JM, Tipnis N, Nurko S. An ANMS-NASPGHAN consensus document on esophageal and antroduodenal manometry in children. Neurogastroenterol Motil. 2018;30(3).

11. Gharibans AA, Hayes TCL, Carson DA, Calder S, Varghese C, Du P, et al. A novel scalable electrode array and system for non-invasively assessing gastric function using flexible electronics. Neurogastroenterol Motil. 2023;35(2):e14418.

12. Calder S, Schamberg G, Varghese C, Waite S, Sebaratnam G, Woodhead JST, et al. An automated artifact detection and rejection system for body surface gastric mapping. Neurogastroenterol Motil. 2022;34(11):e14421.

13. Sebaratnam G, Karulkar N, Calder S, Woodhead JST, Keane C, Carson DA, et al. Standardized system and App for continuous patient symptom logging in gastroduodenal disorders: Design, implementation, and validation. Neurogastroenterol Motil. 2022;34(8):e14331.

14. Schamberg G, Varghese C, Calder S, Waite S, Erickson J, O’Grady G, et al. Revised spectral metrics for body surface measurements of gastric electrophysiology. Neurogastroenterol Motil. 2023;35(3):e14491.

15. Gharibans AA, Calder S, Varghese C, Waite S, Schamberg G, Daker C, et al. Gastric dysfunction in patients with chronic nausea and vomiting syndromes defined by a noninvasive gastric mapping device. Sci Transl Med. 2022;14(663):eabq3544.

16. Wang WJ, Foong D, Calder S, Schamberg G, Varghese C, Tack J, et al. Gastric Alimetry Expands Patient Phenotyping in Gastroduodenal Disorders Compared with Gastric Emptying Scintigraphy. Am J Gastroenterol. 2024;119(2):331–41.

17. Humphrey G, Keane C, Schamberg G, Benitez A, Calder S, Xu B, et al. BODY SURFACE GASTRIC MAPPING DELINEATES SPECIFIC PATIENT PHENOTYPES IN ADOLESCENTS WITH FUNCTIONAL DYSPEPSIA AND GASTROPARESIS. Digestive Disease Week 2024.

18. Xu W, Gharibans AA, Calder S, Schamberg G, Walters A, Jang J, et al. Defining and Phenotyping Gastric Abnormalities in Long-Term Type 1 Diabetes Using a Novel Body Surface Gastric Mapping Device. 2023.

19. Russell AC, Stone AL, Wang A, Walker LS. Development and Validation of a Nausea Severity Scale for Assessment of Nausea in Children with Abdominal Pain-Related Functional Gastrointestinal Disorders. Children (Basel). 2018;5(6).

20. Banks KP, Syed K, Parekh M, McWhorter N. Gastric Emptying Scan. StatPearls. Treasure Island (FL) ineligible companies. Disclosure: Kunzah Syed declares no relevant financial relationships with ineligible companies. Disclosure: Maansi Parekh declares no relevant financial relationships with ineligible companies. Disclosure: Nathan McWhorter declares no relevant financial relationships with ineligible companies. 2024.

21. Chanpong A, Cronin H, Rampling D, Ashworth M, Eaton S, Rybak A, et al. Enhancing the utility of antroduodenal manometry in pediatric intestinal pseudo-obstruction. Neurogastroenterol Motil. 2022;34(5):e14259.

22. Foong D, Calder S, Varghese C, Schamberg G, Xu W, Daker C, et al. Gastric Alimetry((R)) Test Interpretation in Gastroduodenal Disorders: Review and Recommendations. J Clin Med. 2023;12(20).

23. Humphrey G, Schamberg, G., Keane, C., Calder, S., Andrews, C.N., Barsamian, J.M., Bowerman, C., Klienman, S., Benitez, A.J., O’Grady, G., Mousa, H.M., & Gharibans, A.A.. TOWARDS AGE SPECIFIC NORMATIVE VALUES FOR BODY SURFACE GASTRIC MAPPING IN PEDIATRICS. NASPGHAN 2023 2023.

24. Law M, Schamberg G, Gharibans A, Sebaratnam G, Foong D, Varghese C, et al. Short– and long-term reproducibility of body surface gastric mapping using the Gastric Alimetry(R) system. Neurogastroenterol Motil. 2024:e14812.

25. Lin Z, Sarosiek I, Forster J, Damjanov I, Hou Q, McCallum RW. Association of the status of interstitial cells of Cajal and electrogastrogram parameters, gastric emptying and symptoms in patients with gastroparesis. Neurogastroenterol Motil. 2010;22(1):56–61, e10.

26. Angeli TR, Cheng LK, Du P, Wang TH, Bernard CE, Vannucchi MG, et al. Loss of Interstitial Cells of Cajal and Patterns of Gastric Dysrhythmia in Patients With Chronic Unexplained Nausea and Vomiting. Gastroenterology. 2015;149(1):56–66 e5.

27. O’Grady G, Angeli TR, Du P, Lahr C, Lammers W, Windsor JA, et al. Abnormal initiation and conduction of slow-wave activity in gastroparesis, defined by high-resolution electrical mapping. Gastroenterology. 2012;143(3):589–98 e3.

28. Pasricha PJ, Grover M, Yates KP, Abell TL, Koch KL, McCallum RW, et al. Progress in Gastroparesis – A Narrative Review of the Work of the Gastroparesis Clinical Research Consortium. Clin Gastroenterol Hepatol. 2022;20(12):2684–95 e3.

29. Cogliandro RF, Rizzoli G, Bellacosa L, De Giorgio R, Cremon C, Barbara G, et al. Is gastroparesis a gastric disease? Neurogastroenterol Motil. 2019;31(5):e13562.

30. Camilleri M, Malagelada JR. Abnormal intestinal motility in diabetics with the gastroparesis syndrome. Eur J Clin Invest. 1984;14(6):420–7.

31. Grover M, Farrugia G, Lurken MS, Bernard CE, Faussone-Pellegrini MS, Smyrk TC, et al. Cellular changes in diabetic and idiopathic gastroparesis. Gastroenterology. 2011;140(5):1575–85 e8.

32. Huizinga JD, Zarate N, Farrugia G. Physiology, injury, and recovery of interstitial cells of Cajal: basic and clinical science. Gastroenterology. 2009;137(5):1548–56.

33. Kornum DS, Terkelsen AJ, Bertoli D, Klinge MW, Hoyer KL, Kufaishi HHA, et al. Assessment of Gastrointestinal Autonomic Dysfunction: Present and Future Perspectives. J Clin Med. 2021;10(7).

34. Langford JS, Tokita E, Martindale C, Millsap L, Hemp J, Pace LA, et al. Quantitative gastrointestinal function and corresponding symptom profiles in autonomic neuropathy. Front Neurol. 2022;13:1027348.

35. Mehr SE, Barbul A, Shibao CA. Gastrointestinal symptoms in postural tachycardia syndrome: a systematic review. Clin Auton Res. 2018;28(4):411–21.

36. Huang IH, Calder S, Gharibans AA, Schamberg G, Varghese C, Andrews CN, et al. Meal effects on gastric bioelectrical activity utilizing body surface gastric mapping in healthy subjects. Neurogastroenterol Motil. 2024:e14823.

