## Supplementary Material 2 for "Neuropathic Gastroduodenal Disorders can be Diagnosed by Non-Invasive Body Surface Gastric Mapping: a Comparison with Antroduodenal Manometry"

| Supporting Information 2: Patient Demographics | | |
|---|---|---|
| **Variable** | **Description** | **n** |
| **Gender** | Female | 13 |
| | Male | 2 |
| **Age** | Mean | 15.9 ±2.7 years |
| | Range | 10-19 years |
| **BMI** | Average | 20.6 ±3.5 |
| **Diagnosis upon enrollment** | Gastroparesis | 8 |
| | Functional Dyspepsia | 7 |

Supporting Information 2: Patient Demographics
