## Supplementary figures and images for "Neuropathic Gastroduodenal Disorders can be Diagnosed by Non-Invasive Body Surface Gastric Mapping: a Comparison with Antroduodenal Manometry"

### Supplementary Material 3

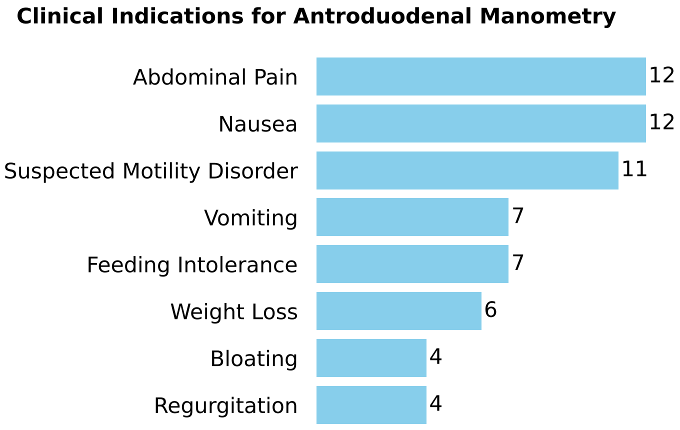
