## Supplementary Material 1 for "Neuropathic Gastroduodenal Disorders can be Diagnosed by Non-Invasive Body Surface Gastric Mapping: a Comparison with Antroduodenal Manometry"

| Supporting Information 1. Normative Reference Intervals for Body Surface Gastric Mapping Metrics in Pediatrics | | | | |
|---|---|---|---|---|
| **Metric** | **Low Reference Value** | **Borderline Low Interval** | **High Reference Value** | **Borderline High Interval** |
| Principal Gastric Frequency (cpm) | 2.65 | 2.6-2.75 | 3.35 | 3.3-3.4 |
| BMI-Adjusted Amplitude (uV) | 22 | 20-25 | 70 | 60-100 |
| Max Postprandial Gastric Alimetry Rhythm Index | 0.25 | 0.19-0.28 | - | - |
| Fed:Fasted Ratio | 1.08 | 1.01-1.12 | - | - |

Supporting Information 1: Normal Pediatric Body Surface Gastric Mapping Parameters
